# CD83 Receptor Unveiled as a Therapeutic Target in Metastatic Breast Cancer through Single-Cell RNA-Seq

**DOI:** 10.1101/2024.01.30.24302004

**Authors:** Saed Sayad, Mark Hiatt, Hazem Mustafa

**Affiliations:** Bioada Lab

## Abstract

**Background:** Metastatic breast cancer is an advanced stage of the disease in which cancer cells have spread to organs beyond the breast, posing a significant clinical challenge, necessitating the development of novel therapeutic approaches. Gaining a profound understanding of the intricacies of metastasis is essential for developing effective interventions. In this study, we investigated the CD83 gene and its receptor as a promising therapeutic target in metastatic breast cancer.

**Method:** We obtained single-cell transcriptomes (*GSE75688*) from the NIH portal website. These transcriptomes encompassed samples from eleven patients (BC01-BC11), which included two lymph node metastases (BC03LN, BC07LN). Our analysis focused on identifying genes with distinct expression patterns between metastatic and primary single cells.

**Results:** In the array of 22 tables (comprising two metastatic and eleven primary tables) featuring the top 800 differentially expressed genes in each, CD83 stood out as the sole significantly upregulated gene consistently shared across all eleven patients at the intersection. CD83 is a protein that plays a crucial role in the immune system, particularly in the context of antigen presentation, immune regulation and immune checkpoint, suggesting its potential involvement in breast cancer progression and metastasis. Based on these findings, we hypothesized that blocking the CD83 receptor on the cell membrane using an antibody could effectively inhibit cell growth in metastatic breast cancer.

**Conclusion:** We propose a novel therapeutic approach by targeting the CD83 receptor on the cell membrane to inhibit cell growth and metastasis in breast cancer. While these findings are promising, further research is needed to validate the efficacy and safety of CD83-targeted therapies. If successful, such treatments could offer improved options for patients with metastatic breast cancer, addressing a critical need for more effective interventions in advanced stages of the disease.

## Introduction

Metastatic breast cancer represents the advanced stage of this daunting disease, marking a critical juncture in its progression. At this stage, cancer cells have spread beyond the breast and nearby lymph nodes to other organs, such as the lungs, liver, bones, and brain, posing significant challenges to effective treatment. Unlike localized breast cancer, metastatic breast cancer requires a comprehensive and nuanced approach due to its systemic nature. Understanding the intricacies of metastasis is pivotal in developing targeted therapies and interventions that can make a meaningful impact in the battle against this formidable adversary. The survival rates for metastatic breast cancer vary based on several factors. In the United States, the 5-year relative survival rate for women with metastatic breast cancer is 30% [1]. Cluster of Differentiation (CD) proteins emerge as key players influencing tumor development, metastasis, and therapeutic responses. CD proteins in breast cancer signify their multifaceted roles in diagnosis [2], prognosis [3], immune modulation [4], targeted therapy [5], metastasis [6], and treatment resistance [7]. Importantly, while some CD proteins may promote metastasis, others may act as tumor suppressors, inhibiting the spread of cancer cells. In this study, we focused on CD83, a cell surface receptor found to be upregulated in single cells extracted from breast cancer patients.

## Data

We downloaded single-cell transcriptomes (*GSE75688*) from the NIH portal website. All single-cell mRNA expression profiles (**Figure 1**) were acquired from eleven patients (BC01-BC11), including two lymph node metastases (BC03LN, BC07LN). We applied a log2 transformation to the dataset prior to analysis.

**Figure 1:**
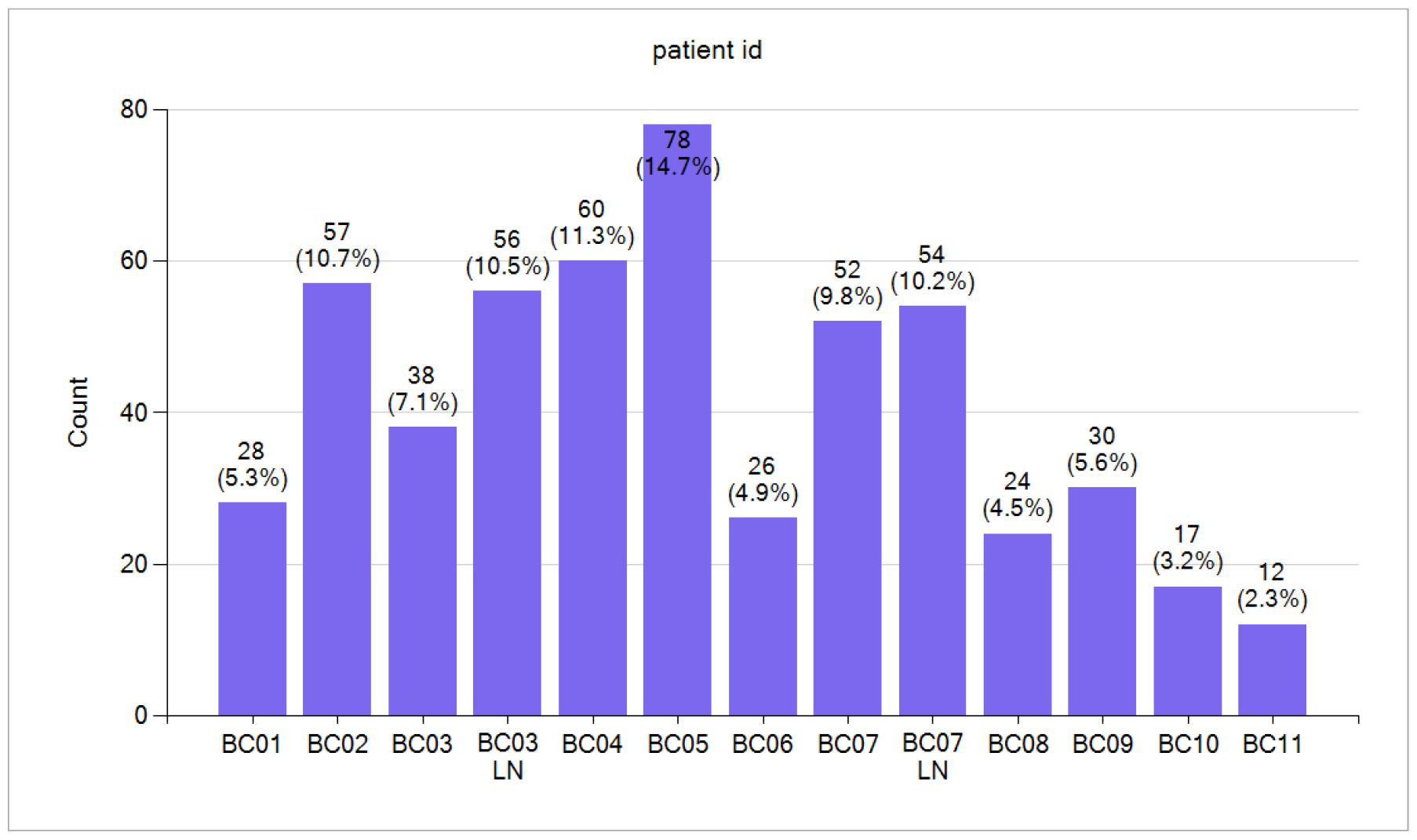
Single-cell count for eleven primary breast cancer patients and two lymph node metastases (LN).

## Data Analysis

In this study, we performed a comprehensive comparative analysis focusing on the single-cell transcriptomes of both primary and metastatic breast cancer. This investigation aimed to unravel the detailed patterns of gene-expression patterns underlying the progression of the disease. With single-cell transcriptomes allowing us to examine individual cells, we could discern subtle variations in gene-expression profiles, gaining insights into the diverse cell populations that contribute to the dynamic nature of breast-cancer progression. The primary tumor serves as the origin point of malignancy, and understanding the transcriptomic alterations occurring at this site is crucial for deciphering the initial stages of cancer development. We went further in our study, looking at metastatic lesions. This gave us a special view of how cancer cells change at the molecular level to spread and grow in other parts of the body.

### Differential Gene Expression – Primary Cells against Metastatic Cells

Using the t-test, we compared the primary breast cancer cells (422 cells from eleven patients) with the metastatic cells from two patients (110 cells) to find differentially expressed genes (**Figure 2**). This statistical comparison provides insight into molecular distinctions between primary and metastatic cancer cell populations, contributing valuable information to our understanding of breast-cancer dynamics. Out of many differentially expressed genes, we selected top ten upregulated and downregulated genes (**Table 1**) that represent a diverse range of biological pathways and functions, but are especially connected to the immune system.

**Table 1:** Top ten upregulated and downregulated genes through primary and metastatic gene-expression analysis.

| Top 10 Differentially Expressed Genes – Primary Cancer Cells against Metastatic Cancer Cells |  |  |  |
| --- | --- | --- | --- |
| Upregulated |  | Downregulated |  |
| Gene Symbol | Gene Name | Gene Symbol | Gene Name |
| *MS4A1* | Membrane Spanning 4-Domains A1 | *S100A6* | S100 Calcium Binding Protein A6 |
| *CD79B* | CD79b Molecule | *S100A16* | S100 Calcium Binding Protein A16 |
| *LRMP* | Inositol 1,4,5-Triphosphate Receptor Associated 2 | *MT-ND3* | Mitochondrially Encoded NADH |
| *CD83* | CD83 Molecule | *MGP* | Matrix Gla Protein |
| *FCRLA* | Fc Receptor Like A | *TCF7L2* | Transcription Factor 7 Like 2 |
| *PAX5* | Paired Box 5 | *CD63* | CD63 Molecule |
| *LY9* | Lymphocyte Antigen 9 | *IFI6* | Interferon Alpha Inducible Protein 6 |
| *CD69* | CD69 Molecule | *PFN2* | Profilin 2 |
| *SNX29P2* | Sorting Nexin 29 Pseudogene 2 | *MT-ND1* | Mitochondrially Encoded NADH |
| *POU2AF1* | POU Class 2 Homeobox Associating Factor 1 | *HK2* | Hexokinase 2 |

**Figure 2:**
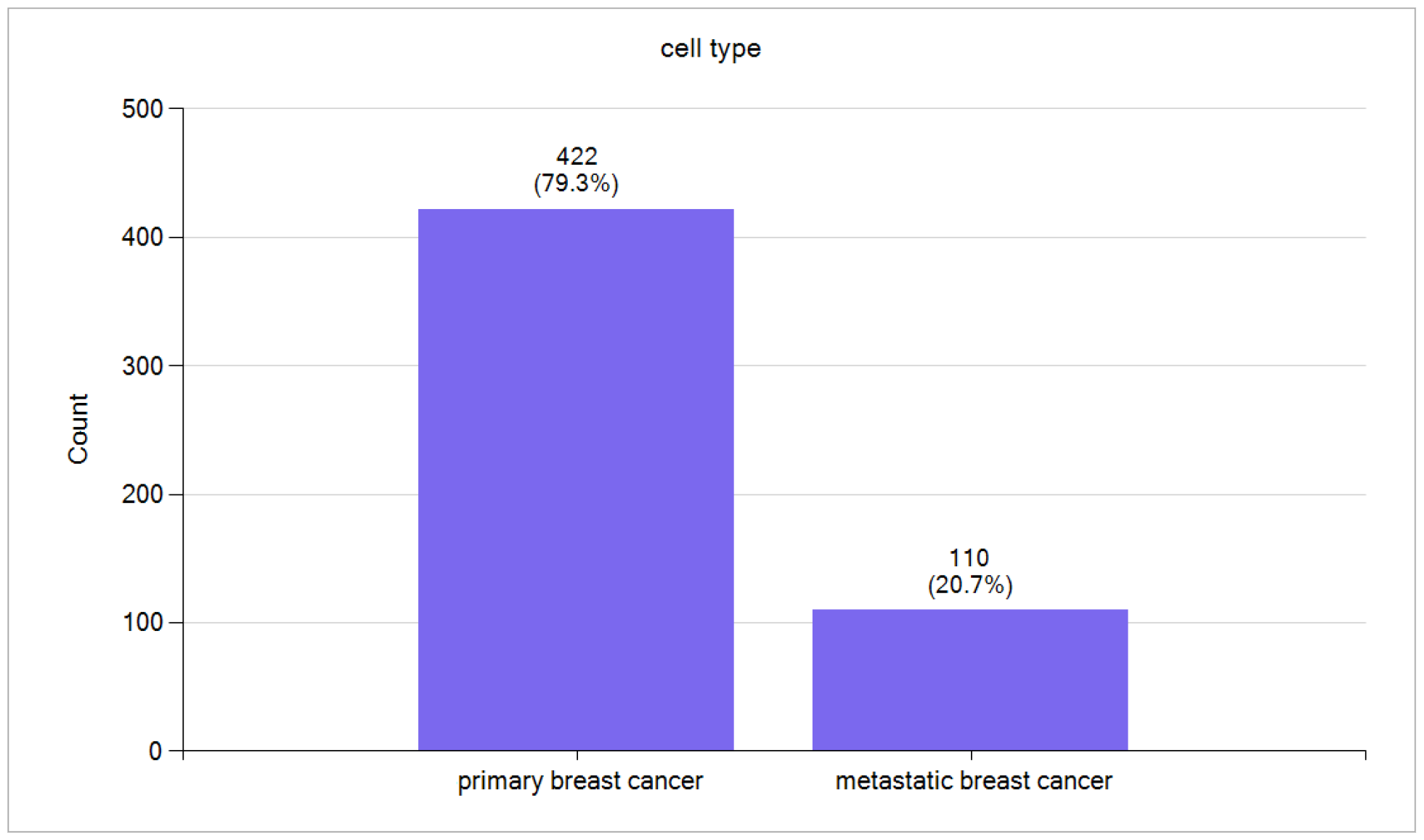
Breast cancer primary single cells from eleven patients and metastatic cells from two patients.

### Differential Gene Expression – Primary Cancer Cells against Primary Cancer Cells

We identified numerous differentially expressed genes upon comparing primary cancer cells with metastatic cancer cells, assuming homogeneity within the primary cancer-cell population. To scrutinize this assumption, we systematically compared primary cancer cells from one patient against those from other patients in eleven separate analyses. Across all paired comparisons, we consistently observed significant upregulation and downregulation of genes. This finding implies that the assumption of all primary cancer cells belonging to a single population is untenable. The differential gene-expression patterns observed in our analyses underscore the importance of considering inter-patient variability in future studies, as it contributes crucial insights into the complexities of cancer biology. The specific case of BC01 against BC02 serves as a representative example (**Figure 3**). We chose the top ten upregulated and downregulated genes (**Table 2**) from a pool of differentially expressed genes, highlighting their significance in diverse biological pathways.

**Table 2:**
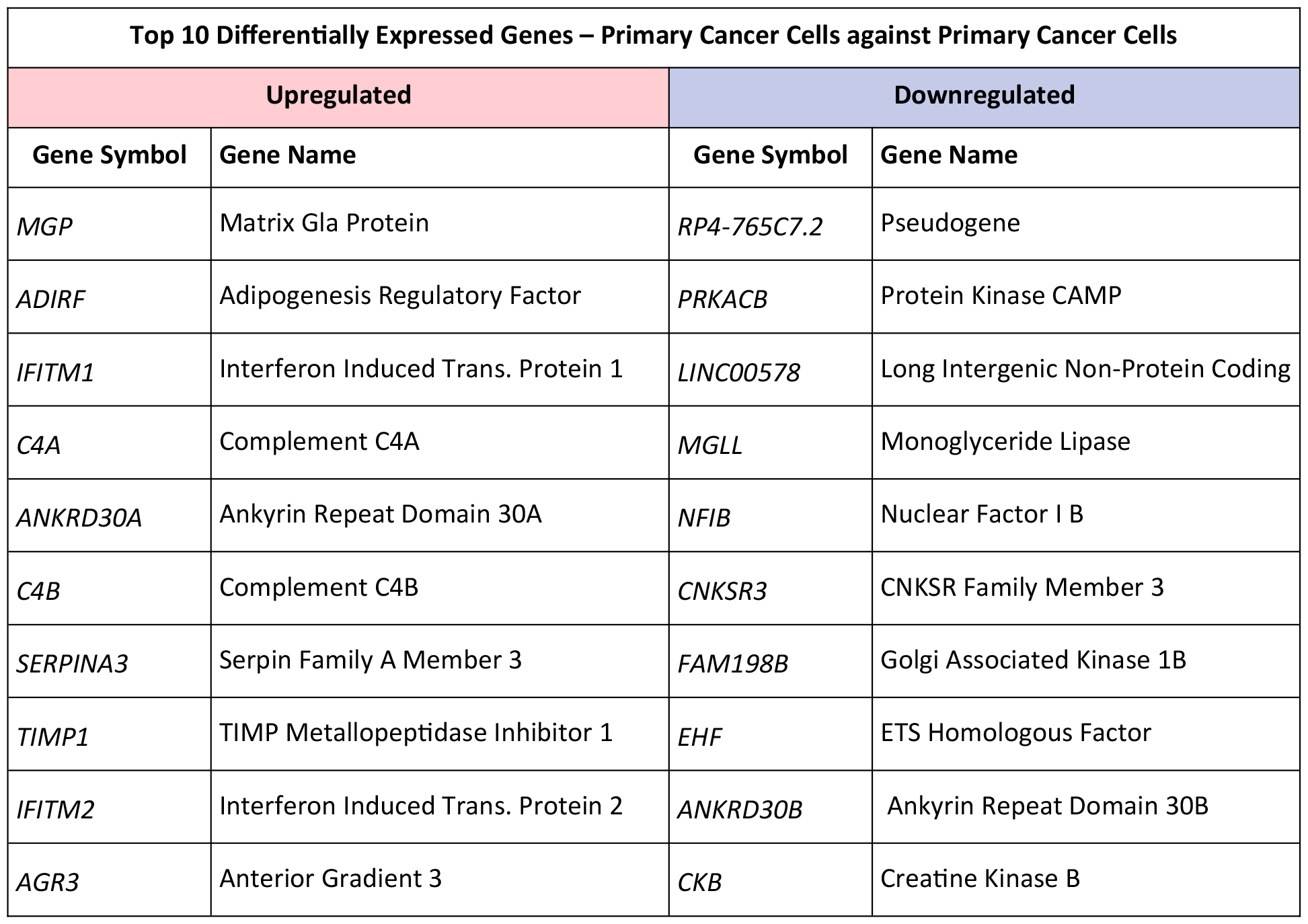
Top ten upregulated and downregulated differentially expressed genes for two patients (BC01 & BC02).

**Figure 3:**
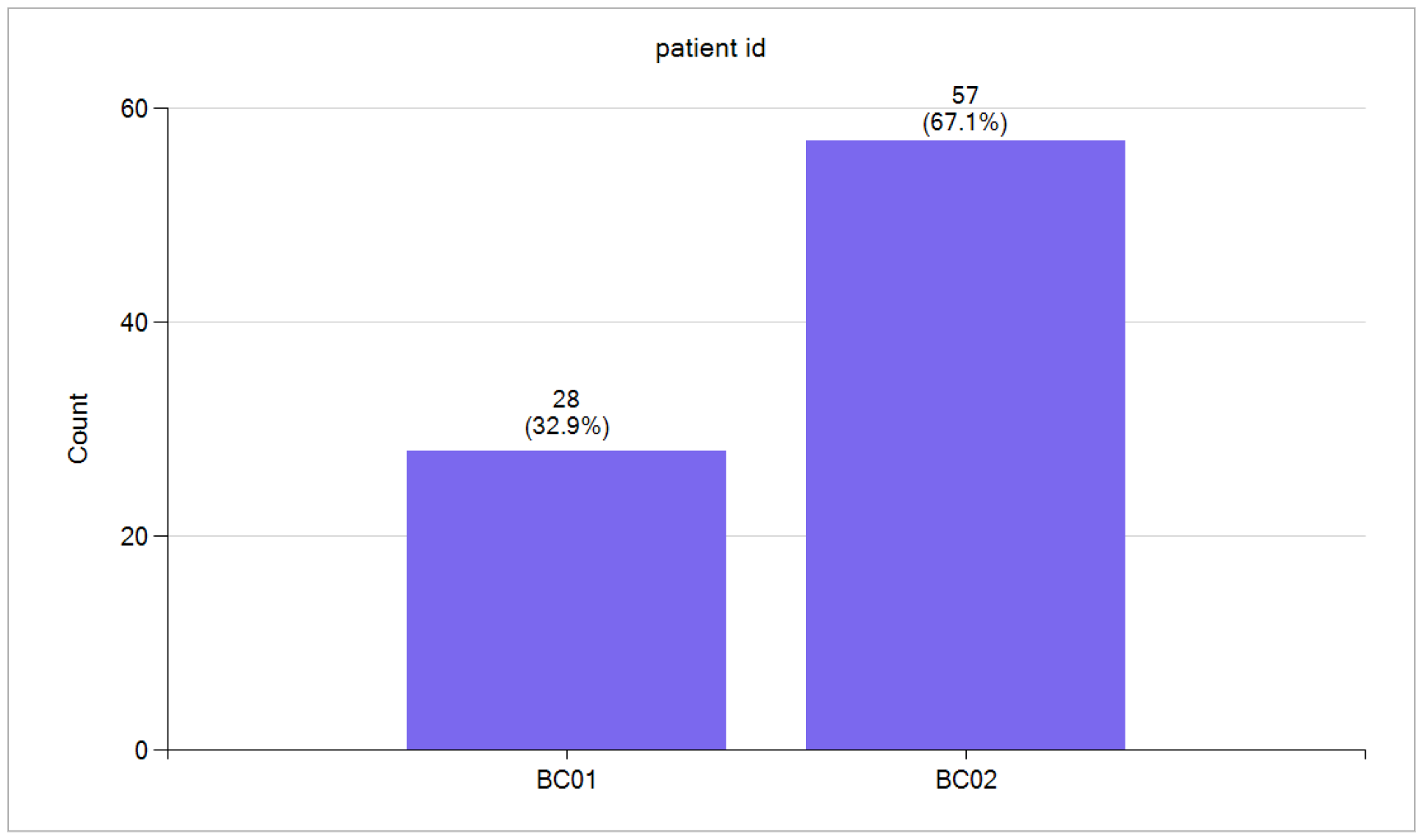
Breast cancer primary single cells from two patients (BC01 and BC02).

### Differential Gene Expression – Comparing Primary Cancer Cells within a Single Patient

We demonstrated that assuming the primary cancer cells from eleven patients originate from a single population is not valid. In our investigation of cancer-cell heterogeneity in a single patient (BC01), we randomly divided the cells into two subsets (**Figure 4**) and employed a t-test to identify differentially expressed genes. However, no significant upregulated or downregulated genes were observed for BC01 or the other ten patients.

**Figure 4:**
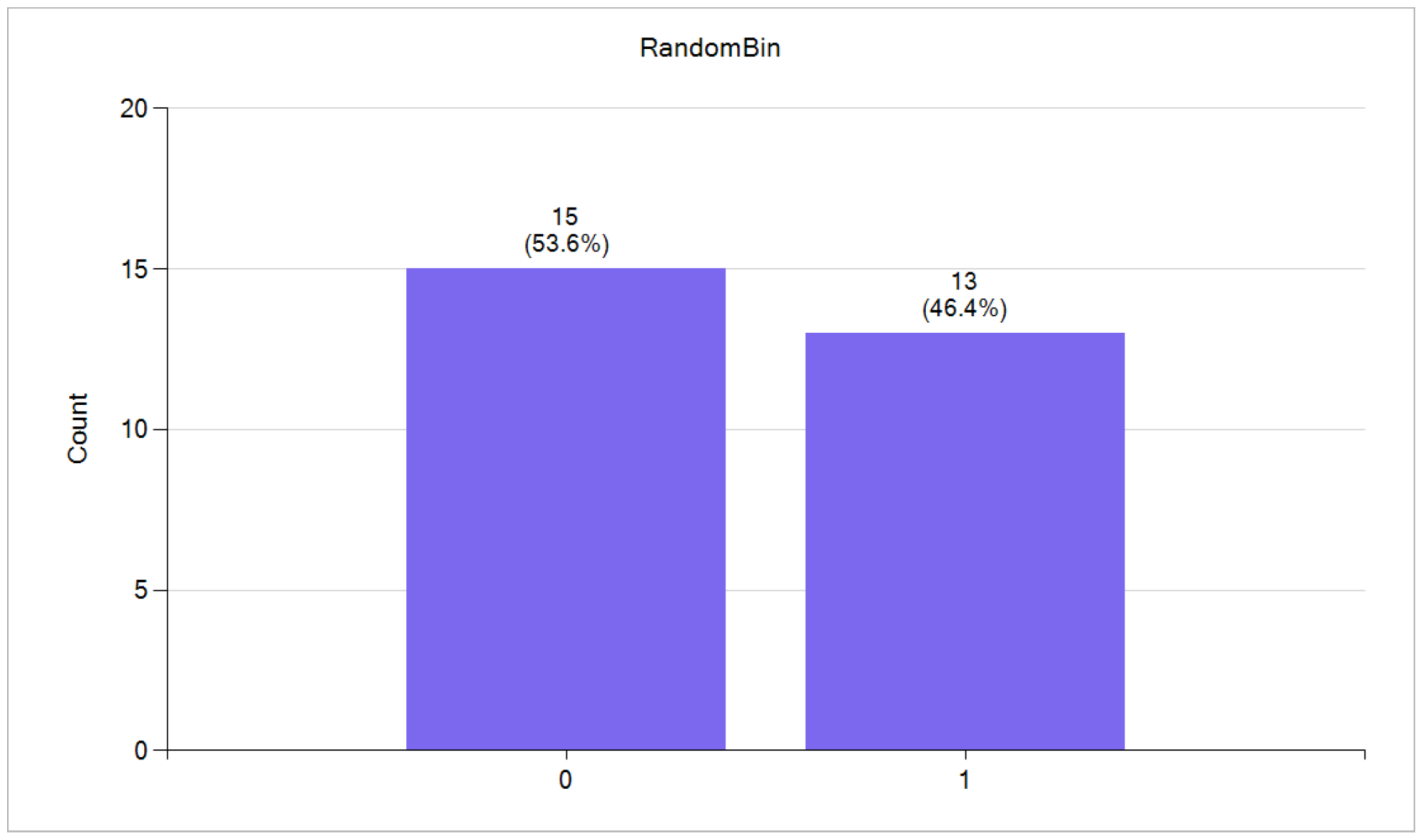
Breast cancer primary single cells from one patient (BC01) divided to two subsets randomly.

### Differential Gene Expression – Metastatic Cancer Cells against Metastatic Cancer Cells

Through a comparison between primary cancer cells and metastatic cancer cells we showed that there are many differentially expressed genes, initially assuming homogeneity within the metastatic cancer cell population. In order to assess this assumption, we systematically compared metastatic cancer cells from one patient with those from another (**Figure 5**). Our findings revealed that the assumption of a unified population among all metastatic cancer cells is untenable. The distinctive patterns of gene expression observed in our analyses underscore the necessity of acknowledging inter-patient variability in future studies, as it provides essential insights into the intricacies of cancer biology. To further elucidate the significance of these findings, we selectively identified the top ten upregulated and downregulated genes (**Table 3**) from a pool of differentially expressed genes, highlighting their significance in diverse biological pathways.

**Table 3:** Top ten upregulated and downregulated differentially expressed genes for BC03LN against BC07LN.

| Top 10 Differentially Expressed Genes – Metastatic Cancer Cells against Metastatic Cancer Cells |  |  |  |
| --- | --- | --- | --- |
| Upregulated |  | Downregulated |  |
| Gene Symbol | Gene Name | Gene Symbol | Gene Name |
| *RPL9P8* | Ribosomal Protein L9 Pseudogene 8 | *RP11-490G8.1* | Pseudogene |
| *RPL9P9* | Ribosomal Protein L9 Pseudogene 8 | *RPL29P11* | Ribosomal Protein L29 Pseudogene 11 |
| *MUCL1* | Mucin Like 1 | *CCDC144B* | Coiled-Coil Domain Containing 144B |
| *CXADR* | CXADR Ig-Like Cell Adhesion Molecule | *KIF23* | Kinesin Family Member 23 |
| *MGST1* | Microsomal Glutathione S-Transferase 1 | *MELK* | Maternal Embryonic Leucine Zipper Kinase |
| *FABP7* | Fatty Acid Binding Protein 7 | *CDK1* | Cyclin Dependent Kinase 1 |
| *CD44* | CD44 Molecule | *RPSAP15* | Ribosomal Protein SA Pseudogene 15 |
| *MARCKS* | Myristoylated Alanine Rich Protein Kinase C Substrate | *PRPSAP2* | Phosphoribosyl Pyrophosphate Synthetase Associated Protein 2 |
| *TSPAN8* | Tetraspanin 8 | *MZB1* | Marginal Zone B And B1 Cell |
| *YBX3* | Y-Box Binding Protein 3 | *USP32P2* | Ubiquitin Specific Peptidase 32<br>Pseudogene 2 |

**Figure 5:**
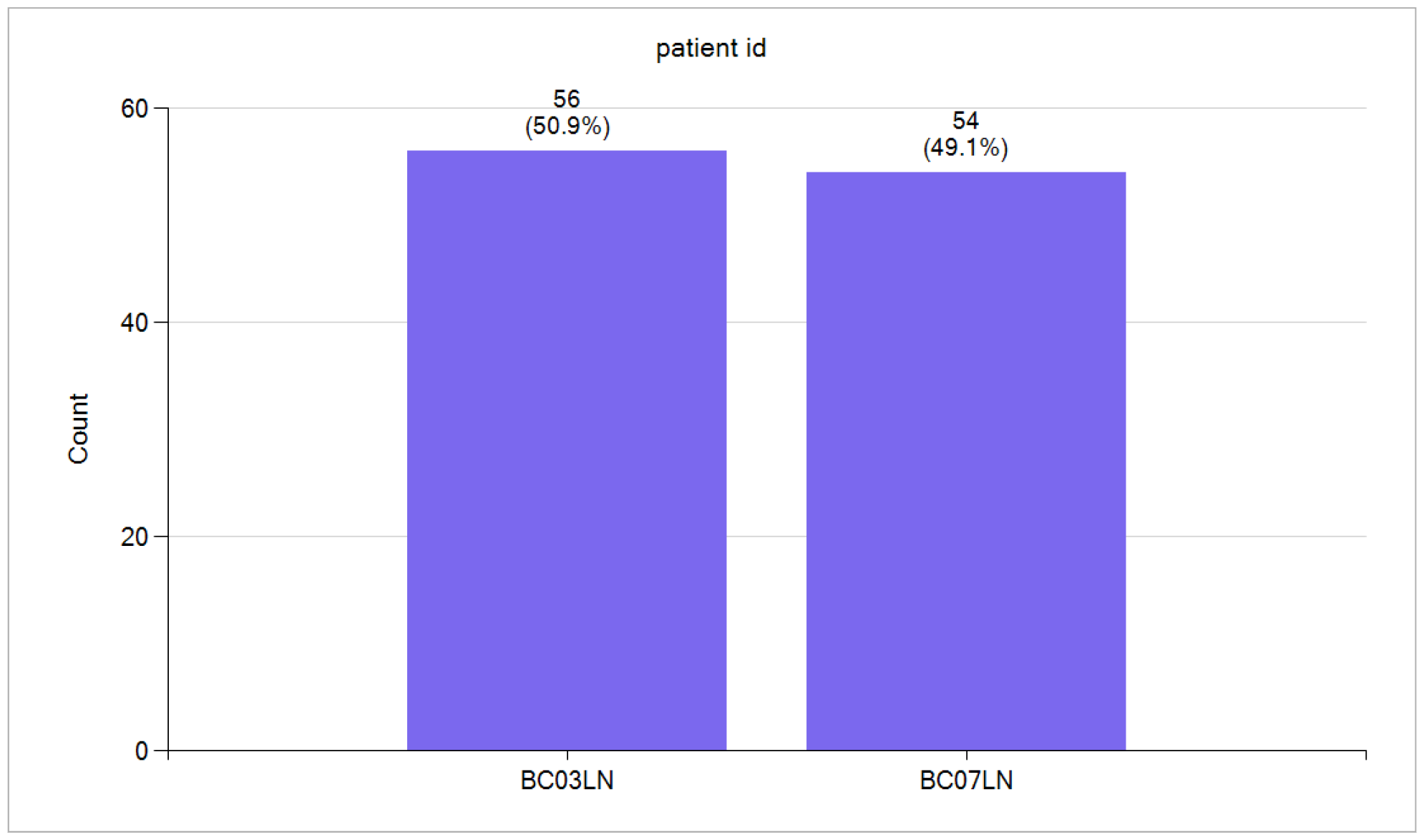
Breast cancer metastatic single cells from two patients (BC03LN and BC07LN).

### Differential Gene Expression – Comparing Metastatic Cancer Cells Within a Single Patient

We demonstrated that assuming the metastatic cancer cells from two patients originate from a single population is not valid. In our investigation of cancer cell heterogeneity in a single patient (BC03LN), we randomly divided the cells into two subsets (**Figure 6**) and employed a t-test to identify differentially expressed genes, but no significant upregulated or downregulated genes were observed. We repeated the same analysis for the second sample (BC07LN) and did not observe any significant upregulated or downregulated genes.

**Figure 6:**
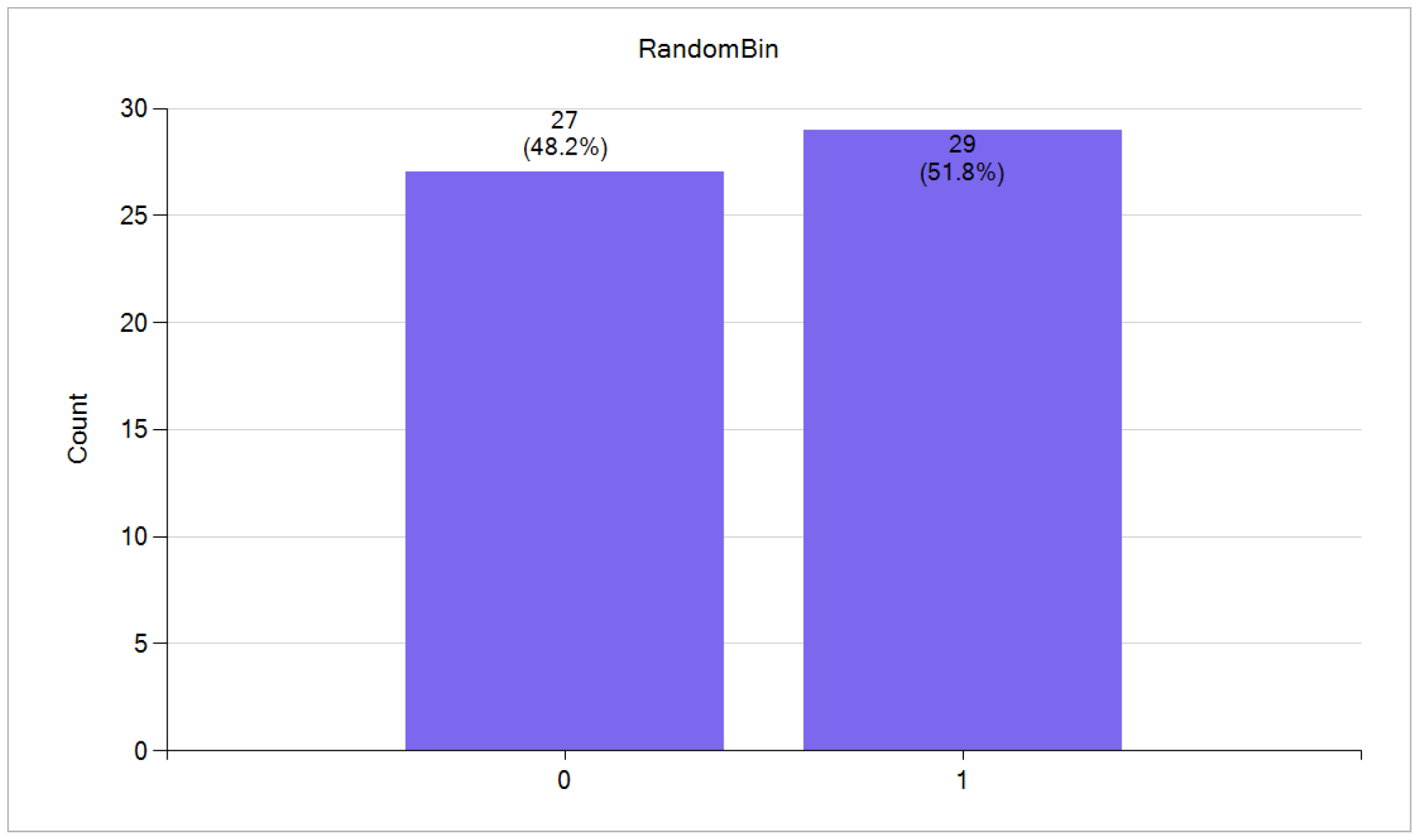
Breast cancer metastatic single cells from one patient (BC03LN) divided to two subsets randomly.

### Differential Gene Expression – Metastatic Cells against Primary Cells – Patient by Patient

Our findings revealed that the assumption of a unified population among all primary or metastatic cancer cells is untenable. We approached the analysis of each patient’s cancer cells by treating them as independent samples. Utilizing a statistical analysis, we systematically compared metastatic cancer cells from each patient against all primary cancer cells (patient by patient). Subsequently, we identified and retained the top 800 upregulated and downregulated genes for each test. In **Table 4**, we presented the top three upregulated and downregulated genes. Notably, the RP4-765C7.2 gene appeared in twelve out of the 22 top-three upregulated genes, while the IFI6 gene was featured in five out of 22 top-three downregulated genes. RP4-765C7.2 is a pseudogene with unknown functionality and IFI6 is one of the many genes induced by interferon [8]. We attempted to identify common genes among 22 tests from the initially saved set of 800 genes for each test. To our surprise, we discovered only one gene that was consistently present across all tests: *CD83*.

**Table 4:**
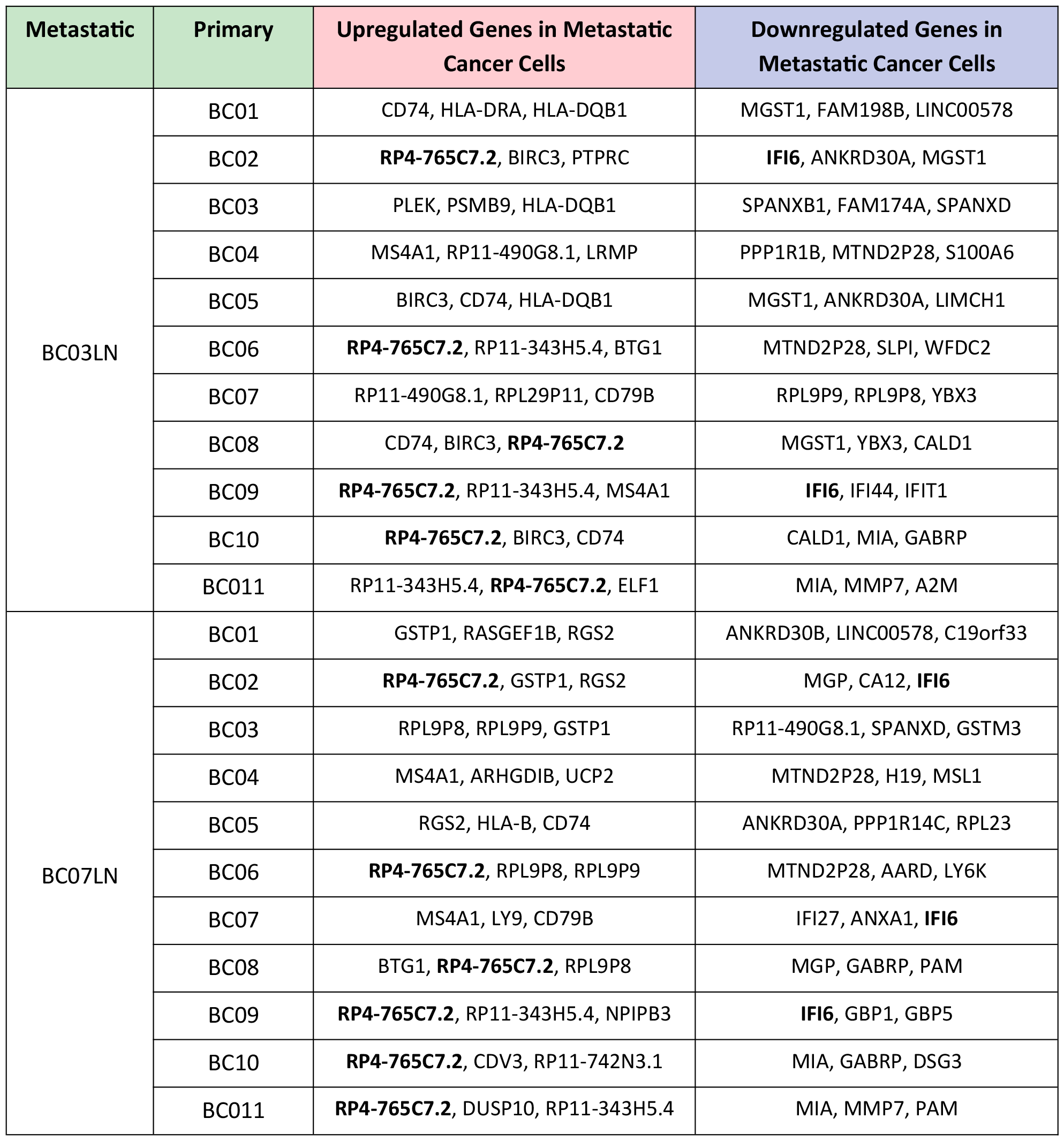
Top three upregulated and downregulated genes by comparing metastatic cells against primary cells.

The CD83 gene is located on human chromosome 6p23. In humans, a promoter 261 bp upstream consists of five NF-κB and three interferon regulatory factor binding sites, reflecting the involvement of CD83 in inflammation [9]. The CD83 protein is a transmembrane glycoprotein that plays pivotal roles in the immune system, particularly in the regulation of T-cell function and antigen presentation. Primarily expressed on the surface of mature dendritic cells (DCs), CD83 serves as a maturation marker, indicating the readiness of DCs to initiate immune responses. Significantly, the CD83 protein emerges as a potential Immune checkpoint [10]. Immune checkpoints are molecules or receptors that play a pivotal role in modulating the immune response, ensuring a delicate balance between activation and inhibition to prevent excessive immune reactions or autoimmunity.

## Discussion

We investigated the complex landscape of metastatic breast cancer, with a particular focus on understanding the significance of the CD83 gene. We conducted a thorough comparative analysis of single-cell transcriptomes derived from both primary and metastatic breast cancer cells. Contrary to the assumption of uniformity within these cell populations, this study revealed substantial variability among patients, challenging the notion of homogeneity. The molecular differences identified between primary and metastatic breast cancer cells provide a deeper understanding of the disease dynamics. The recognition of this heterogeneity emphasizes the necessity for a nuanced comprehension of cancer biology, considering the diverse gene-expression patterns that contribute to the intricacies of the illness. The findings not only underscore the importance of acknowledging heterogeneity, but also pinpoint specific genes, including RP4-765C7.2, IFI6, and CD83, which are shared across diverse patient samples. However, CD83 stood out as the sole significantly upregulated gene consistently shared across all eleven patients at the intersection.

The multifaceted functions of CD83 contribute to the orchestration of immune responses, making this gene a key player in maintaining immune homeostasis and regulating adaptive immune reactions. The role of CD83 in inflammation and its expression on various immune cells position it as a potential target for further exploration in developing targeted therapies. Moreover, recognizing CD83 as an immunity checkpoint adds a layer of complexity and hope to its significance in breast cancer, emphasizing its potential as a key factor in immune modulation and promising avenue for therapeutic interventions. Our study contributes valuable insights that could inform the development of targeted therapies and interventions, offering new approaches in the battle against metastatic breast cancer.

## Summary

This study focused on metastatic breast cancer, an advanced stage in which cancer cells have spread beyond the breast, posing significant treatment challenges. We analyzed single-cell transcriptomes (primary and metastatic) from eleven patients, identifying CD83 as consistently upregulated in metastatic cells. We thus propose blocking the CD83 receptor with an antibody as a novel means to inhibit metastatic breast cancer growth, and emphasize the need for further research to validate the efficacy and safety of this approach to improved treatment.

## Data Availability

https://www.ncbi.nlm.nih.gov/geo/query/acc.cgi?acc=GSE75688

